# ASSESSMENT OF KNOWLEDGE AND DISPOSAL PRACTICES OF SPENT AND BROKEN ENERGY-SAVING BULBS AMONG HOUSEHOLDS IN MTENDERE COMPOUND ZAMBIA

**DOI:** 10.64898/2026.04.02.26349820

**Authors:** Moffat Maselechi

## Abstract

The widespread adoption of energy-saving bulbs like light-emitting diodes and compact fluorescent lamps in Zambia has raised significant environmental and public health issues because some of these bulbs contain dangerous materials like mercury. This study sought to evaluate households’ understanding and disposal practices of used and damaged energy-saving bulbs in Lusaka, Zambia’s Mtendere Compound. A cross-sectional design was used, with structured questionnaires distributed to a randomly chosen sample of households. The research showed that, although most participants were aware of the energy efficiency advantages of these bulbs, they had little understanding of their possible health risks and safe disposal procedures. The majority of households reported throwing away broken and used bulbs with their regular household trash, while only a small percentage followed the suggested disposal procedures. Environmental contamination and heightened health risks are exacerbated by a lack of awareness and inadequate municipal waste management systems for hazardous household waste. The research advocates for improved public education initiatives, the creation of specific collection sites for dangerous waste, and the formulation of explicit national regulations and policies for the handling of discarded and damaged energy-saving bulbs. In rapidly urbanizing areas like Mtendere, tackling these issues is essential for protecting public health and advancing environmental sustainability.

## Introduction

The increasing global imperative for energy efficiency and environmental sustainability has led to a widespread adoption of energy-saving bulbs, particularly Compact Fluorescent Lamps (CFLs) and, more recently, Light Emitting Diodes (LEDs), in residential settings worldwide. (1) These technologies offer significant advantages over traditional incandescent bulbs, primarily in terms of reduced energy consumption and extended lifespan, contributing to lower electricity bills and a smaller carbon footprint. (2) However, the benefits of these energy-efficient lighting solutions are accompanied by a critical environmental and public health concern: the proper management of spent and broken bulbs. (3) This concern is particularly pertinent for CFLs, which contain varying amounts of mercury, a potent neurotoxin that can be released into the environment if these bulbs are improperly disposed of (Environmental Health Perspectives). (4) While LEDs generally do not contain mercury, they do contain other hazardous materials such as lead, arsenic, and various rare earth elements, which also pose environmental risks upon improper disposal. (5)

In many developing countries, including Zambia, the transition to energy-saving bulbs has been actively promoted through government initiatives and public awareness campaigns aimed at mitigating energy crises and fostering sustainable development. (6) Mtendere Compound, a high-density residential area in Lusaka, Zambia, exemplifies a community where such transitions are underway. (7) The socio-economic characteristics of such compounds often present unique challenges to environmental management, including limited access to formal waste disposal infrastructure, prevalent informal waste collection practices, and potentially low levels of awareness regarding the hazards associated with specific waste streams. (8) The improper disposal of spent and broken energy-saving bulbs in such contexts can lead to the contamination of soil and water bodies, posing risks to human health through direct exposure or bioaccumulation in the food chain. (9) For instance, mercury exposure, even at low levels, can lead to neurological damage, developmental problems in children, and kidney dysfunction. (10)

Despite the growing adoption of energy-saving bulbs, there remains a significant knowledge gap concerning household awareness of the environmental and health risks associated with their components, as well as the appropriate disposal practices. (11) This gap is particularly pronounced in communities like Mtendere Compound, where formal waste management systems may not adequately address hazardous waste streams. Previous studies in various regions have highlighted a general lack of public awareness regarding the mercury content in CFLs and the proper disposal protocols, often resulting in these bulbs being discarded with general household waste (Resources, Conservation and Recycling). (12) This practice not only negates some of the environmental benefits of using energy-saving bulbs but also creates new environmental and public health challenges. (13)

This manuscript aims to address this critical knowledge gap by conducting a comprehensive assessment of the knowledge and disposal practices of spent and broken energy-saving bulbs among households in Mtendere Compound, Zambia. (14) The study will investigate the level of awareness among residents regarding the composition of these bulbs, the associated health and environmental risks, and the recommended disposal methods. (15) Furthermore, it will explore the current disposal practices employed by households, identify the factors influencing these practices, and assess the accessibility and effectiveness of existing waste management infrastructure for hazardous waste. (16)

The findings of this research are expected to provide valuable insights into the current state of household knowledge and practices, thereby informing the development of targeted public awareness campaigns, improved waste management policies, and sustainable disposal solutions tailored to the specific needs of communities in Zambia and similar developing regions. (17) Ultimately, this study seeks to contribute to the broader goal of promoting environmentally responsible consumption and disposal patterns, ensuring that the benefits of energy efficiency are realized without compromising public health or environmental integrity. (18)

### Health Effects of Mercury Exposure

Poisoning due to mercury exposure is either through ingestion, inhalation or absorption and leads to some adverse health effects. (19) The United States Environmental Protection Agency (UNEPA) identified mercury as one of the 12 toxic pollutants globally. Mercury becomes too toxic if a person is exposed to more than 0.1mg/kg per day. (20) Once released in the atmosphere, mercury accumulates either in soil, air, or water and is then transformed into a more toxic organic form (Khatoon-Abadi, 2008). (21) This includes mercury-coated bulbs that, if improperly disposed, could affect people. (22)

Random mercury disposal is toxic to soil and affects microbial biomass and enzyme activities. Results from soil samples indicated an increase in the levels of mercury, from 0.5μmol/g to 10μmol/g of dried soil. (23) In French Guyana tropical soil microcosms were spiked with mercury and incubated at 28°C for a month and the results demonstrated that in tropical soil, mercury affects soil microbial communities (Harris-Hellal, 2008). (24) The effects of soil contamination due to mercury exposure indicated a reduced size of bacteria and protozoa due to long-term exposure in Copenhagen, Denmark (Muller, Westergaard, Christensen & Sorensen, 2001). (25) Similar conditions of mercury toxicity could be affecting the environment in Mtendere compound due to mercury toxicity, a result of improper disposal of the substance. Once water is contaminated with mercury, marine life is endangered. (26) Overall antioxidant depletion was verified in fish brains collected at the mercury-contaminated stations at the Ria-de Aveiro, Portugal (Mieiro, Ahmad, Pereira, Duarte & Pacheco, 2010). (27) An abandoned mercury mine proved to have an effect on fish, rice, ambient air, and drinking water in a study done to assess human exposure levels and environmental mercury contamination at Honda Bay (Maramba, Reyes, Francisco-Rivera, Panganiban, Dioquino, Dando, Timbang, Akagi, Eguchi & Fuchigami, 2006). (28) The results of the study showed that total mercury contamination of surface water also exceeded total mercury permissible exposure standards (Maramba et al., 2006). (29) Poor handling of mercury-coated objects could endanger animal life and human beings in the Mtendere compound as suggested by studies in Portugal (Mieiro et al., 2010) and Honda Bay (Maramba et al., 2006). (30)

Wang, Shi & Wei (2002) investigated the accumulation and transformation of mercury in soil after it had been deposited in the soil. (31) The results of the study indicated a positive correlation between atmospheric mercury concentration and the content of mercury in soil, thereby proving the toxicity of mercury if proper methods of disposal are not practiced. (32)

Mercury toxicity was discovered in exposed wild birds, mammals and fish in a study conducted in Northern Canada. (33) The purpose of the study was to investigate the effects of environmental methyl-mercury, and the results revealed that exposure to methyl-mercury by mammals, wild birds, and fish led to behavioral, neurochemical, hormonal, and reproductive changes (Schuehammer, Meyer, Sandheinrich & Murray, 2007). (34) In the state of Michigan, USA, two interns were exposed to mercury after spilling a bottle containing mercury on the floor. (35) That led to clinical intoxication, with one intern complaining of insomnia, mild agitation, and tremulousness (Richard & Stephanie, 2004). (36)

It is evident that exposure to mercury, either in water, air or soil could lead to some health effects to human beings and marine life. (EPHC, 2009). (37)

There seems to be similarities in South Africa and the rest of the world regarding poor handling of mercury. (38) In KwaZulu Natal, mercury contamination in the vicinity of a mercury-processing plant was evident. (39) Mercury was discharged into the Mngceweni River which supplies water to the Inanda dam. A study conducted in the Inanda area revealed that 50% of fish samples and 17% of hair samples taken from villagers had mercury concentration levels that exceeded guidelines levels of the World Health Organization (Papu-Zamxaka, Mathee, Harpham, Barnes, Röllin, Lyons, Jordaan & Cloete, 2010). (40) The same problem of exposure to contaminated water was reported among people consuming fish from the Umgeni River near Cato Ridge (Papu-Zamxaka et al., 2010). (41)

Coal mining in South Africa contributes towards increased levels of mercury toxicity. (42) The country is the second-highest mercury emitter in the world. (43) Most of the emissions arise from coal-fired power stations during electricity generation (Dabrowski, Ashton, Murray, and Leaner & Mason 2008). (44) Utility power generator, ESKOM, has thirteen coal-fired power stations that emit large quantities of mercury into the atmosphere, thereby making the exposed environment toxic (Dabrowski & Mason, 2010). (44) Most coal users who took part in a study by Oosthuizen, John & Somerset (2010) indicated increased levels of mercury in their bodies after urine samples were taken and analyzed. (45) Most of them were exposed to mercury either through the inhalation of mercury dust or after consuming contaminated fish or drinking water from polluted water. (46) The source of mercury was a gold mine that was situated next to their residential area that emitted some mercury particles during production (Oosthuizen, John & Somerset, 2010). (47)

### Legislative Control

According to the United States Environmental Protection Agency (2009), mercury coated bulbs contain mercury, and fused bulbs are regarded as hazardous waste under federal and state regulation. (48) However, in some States, hazardous waste bulbs may be managed as universal waste. (49) It is therefore, the responsibility of the generator of mercury coated bulbs to determine whether the bulbs are hazardous waste and to ensure that fused bulbs are in accordance with federal and States regulations. (50)

In Australia, a flourocycle which is a scheme aimed at increasing recycling of mercury coated bulbs was established in 2009 in collaboration with government, industries and the Environment Protection and Heritage Council. (51) Waste disposal and handling is primarily a state and local government responsibility in Australia. (52) Landfill disposal of large amounts of mercury coated bulbs such as those generated by businesses, institutions, or councils is forbidden in the state (Environment Protection and Heritage Council, 2009). (53) In Minnesota, any mercury containing products should not be disposed in landfill, but must be recycled. (54) Their law regulates all mercury containing products including mercury coated bulbs, mercury vapor and metal halide lamps (Minnesota Pollution Agency, 2007). (55)

To ensure that no health or environmental hazards from mercury containing products during their entire life cycle, the European Union has issued a few directives which reduce or ban harmful substances and on the other hand regulate the disposal methods of mercury containing products. (56) The directives regulate the use of certain chemical substances and the disposal of mercury coated bulbs. (57) In Germany, for example, the European directive is implemented by the national law regulating the introduction, collection and environmentally friendly disposal of electric and electronic equipment (Osram, 2013). (58)

Florida laws forbid the disposal of mercury coated bulbs at solid waste incineration facilities. (59) Florida has waste-to-energy facilities that incinerate solid waste to produce energy for the State, and people are not allowed to dispose mercury coated bulbs at these facilities. (60) Besides forbidding disposal at incineration facilities, Florida law also forbid people from throwing mercury coated bulbs away at any landfills or municipal solid waste disposal facilities in the State. (61) The law directs landfill operators to assist enforce disposal prohibition. (62) Florida has permitted reclamation facilities where people can take mercury coated bulbs for recycling (Chinn, 2013). (63) Environment Canada uses regulatory tools under the Canadian Environmental Protection Act (CEPA), 1999 and the Fisheries Act to manage toxic substances such as mercury. (64) Mercury has been deemed a toxic substance under CEPA and is listed on schedule 1 of the Act. (65) There are requirements under CEPA for the management of mercury relating to the chlor-alkali industry, the movement of hazardous waste, environmental emergencies and emissions from various sectors in the National Pollutant Release Inventory. (66) Environment Canada is also involved in the research, development and implementation of a non-regulatory initiative to help reduce and manage releases of mercury due to human activity. (67) The provinces and territories of Canada have legislation, regulations and guidelines for mercury level in liquid effluent, drinking water and emissions from industrial sources. (68) There are also several non-government organizations in the country dedicated to environmental protection that incorporate mercury management strategies into various initiatives (Canada-Ontario Agreement Respecting the great Lakes Basin Ecosystem, 2005). (69)

According to Venter & Van der Walt (2008), the use of mercury coated bulbs becomes increasingly widespread in South Africa so also increases the concerns relating to their mercury content and the associated hazards. (70) The current legislative framework governs large scale users of mercury coated bulbs, but legislation pertaining to industry regarding industry-specific waste is vague and as yet unresolved. Similarly, the legal requirements of residential consumers relating to any hazardous waste is inferred and practically non-existent. (71) Furthermore, waste separation and recycling are not generally practiced and hence unfamiliar concepts to most South Africans. (72)

### Knowledge and practices

Most people who use mercury coated bulbs are unaware that each bulb contains between 5 and 30mg of mercury (Eco-South Travel, 2013). (73) Mercury coated bulbs were found disposed improperly in the metropolitan area on Minas Gerais, Brazil. (74) A study conducted there found out that most people from the general public to those in commercial and health sectors did not know how to dispose end-of-life bulbs coated with mercury (Claudio & Hurbert, 2015). (75) Lack of knowledge regarding the handling of mercury led to about 186 000 kg of mercury being deliberately discharged into the main drainage system of the Maramo Lagoon, Northern Adriatic Sea in Italy. (76) Most of the mercury discharged was from industrial activities. Another secondary discharge experienced at the lagoon was from Idrija in Slovenia (Stefano, Alessandro, Raffaella, Sergio & Cinzia, 2009). (77) The parks and recreation department warned residents near South Dallas playground to be on the lookout for symptoms of mercury exposure after mercury coated bulbs were illegally disposed at a playground and a vacant lot across the street. (78) Nearly 1000 of smashed remains of mercury coated bulbs were found at the Peary play lot in the 2800 block of Peary Avenue, near Malcolm X Boulevard. (79) Reports indicate that the toxic shards were disposed sometime in the evening and were later discovered by parks and recreation service, local police, storm water management, the environmental quality office and local fire rescue, all of whom were called on the scene to investigate. The park was closed due to this improper practice of disposal of mercury coated bulbs (Davis, 2013). (80) Mercury coated bulbs are one of the key measures for addressing electric power shortages and climate change mitigation, and mercury coated bulbs are expected to dominate the lighting in China. (81) Although these bulbs are used in large quantities, residents and industries using liquid mercury are practicing improper disposal methods. (82) It is estimated that spent mercury coated bulbs accounts for approximately 20% of mercury input in China (Hu & Cheng, 2012). (83) According to Sumanapala (2013), Sri Lanka’s public lacks knowledge on proper disposal of mercury containing products such as mercury coated bulbs. (84) Sri Lanka’s health ministry spokesperson Dharma Wanninayake indicated that the ministry was aware of the health implications of the appliances containing mercury. (85) The ministry was mainly concerned about the disposal of the bulbs as people had no knowledge about it. (86) It was therefore, the ministry’s responsibility to ensure that the public receive education or awareness programmes on proper practice of disposal of mercury coated bulbs. (87)

A Vice Mayor in Burlingame was concerned about lack of information with regard to proper disposal of mercury coated bulbs. (88) Even though the State made disposing of mercury coated bulbs in household waste illegal in 2006, community awareness programmes about disposal methods was still a problem. (89) Lack of education to the public about the harmful impact of not recycling the bulbs, contributes to improper practice by the public of disposing mercury coated bulbs in general household waste. (90) The problem was also with the manufacturers of mercury coated bulbs who were providing contradictory information to consumers about proper disposal of mercury coated bulbs (Haughey, 2016). (91)

A study was conducted by Hedge and Hunt (2010) to assess young consumers-college student’s knowledge of compact fluorescent lamps regarding the sustainability, energy usage and their willingness to use it in their home. (92) As energy and sustainability issues become critical, the governments around the world have passed regulations to phase out the inefficient incandescent lamps in favor of more efficient light sources such as mercury coated bulbs. (93) The study surveyed 168 college student’s knowledge about sustainability and energy efficient regarding mercury coated bulbs. (94) Results indicated that 65% of the students believes that mercury coated bulbs does not contain mercury and 77% dispose them in general household waste. (95)

### Disposal Option

In Australia, landfill disposal is prohibited; an alternative to landfill disposal is taking fused mercury coated bulbs to specialized recyclers who are able to safely recover the mercury, glass, phosphor and aluminum contained in the bulbs. (96) Recovered mercury can be re-used in generating new mercury coated bulbs. (97) Several States in Australia also have household chemical collection programmes or drop off center that accept domestic quantities of mercury coated bulbs for recycling (Environment Protection and Heritage Council, 2009). (98)

Mercury containing Lamp Waste Management (2012), recommend that when fused mercury coated bulbs are removed and replaced with new bulbs, the used bulbs should be properly packaged in the cardboard boxes that contained the replacement bulbs. The boxes containing the hazardous mercury coated bulbs waste must be labeled. A safe disposal area should be designated and identified to ensure that the bulbs are not accidentally broken or crushed before they are sent to a disposal facility. The disposal area must be well ventilated, with fire extinguisher and appropriate emergency response equipment in case of accidental emission. Each box of fused bulbs must be stored in a manner that will prevent breakage or damage to the bulbs.

The United States Environmental Protection Agency (2010) further recommended the following steps for cleaning broken mercury coated bulbs:

### Ventilate the room

➢ Have people and pets leave the room, and don’t let anyone walk through the breakage area on their way out.
➢ Open a window and leave the room for 15 minutes or more.
➢ Switch off the air conditioning system.

### Cleaning steps for hard surfaces

➢ Carefully scoop up glass pieces and powder using stiff paper or cardboard and place them in a glass jar with metal lid or in a sealed plastic bag.
➢ Use sticky tape to pick up any remaining small fragments and powder.
➢ Wipe the area clean with damp paper towels or disposable wet wipes. Place towels in the glass jar or plastic bag.
➢ Do not use a vacuum or broom to clean the broken bulb on hard surface.

### Cleaning steps for carpeting or rug

➢ Carefully pick up glass fragments and place them in a glass jar with metal lid or sealed plastic bag.
➢ Use sticky tape to pick up any remaining small glass fragments and powder.
➢ If vacuum is needed after all visible materials are removed, vacuum the area where the bulb was broken.
➢ Remove the vacuum bag and put the bag or vacuum debris in a sealed plastic bag.

### Cleaning steps for clothing, bedding and other soft materials

- If clothing or bedding materials come in direct contact with mercury powder from broken mercury coated bulb, the clothing or bedding should be thrown away. Do not wash such clothing or bedding because mercury fragments in the clothing may contaminate the machine and pollute sewage.
- You can, however, wash clothing or bedding or other materials that have been exposed to mercury vapor, such as the clothing you are wearing when you clean the broken mercury coated bulb.
- If shoes come into direct contact with broken mercury coated bulb, wipe them off with damp paper towels or disposable wet wipes.
- Place the towels or wipes in a glass jar or plastic bag for disposal.

### Disposal of cleaning material

➢ Immediately place all cleaning materials outdoors in a waste container or protected area for the next normal waste collection.
➢ Wash hands after disposing the jars or plastic bags containing cleaning materials.
➢ Check with the local or state government about disposal requirements in your area.

### The Kenya Situation

In its efforts to manage mercury pollution, Kenya set standards on environmental media. The Constitution of Kenya (Aug 2010) and the Environmental Management and Coordination Act (EMCA) of 1999 both underscore the right of every person to a clean and healthy environment and commits each and every one of us to safeguard and enhance the environment. EMCA Waste Management regulations (Legal Notice No121) Fourth Schedule (Regulation 16) defines mercury wastes considered hazardous. However, regulations pertaining to disposal of mercury and mercury-containing products have not been formulated. Amina Ibakari, (2012)

### In Zambia

According to the Government of the republic of Zambia (GRZ) (2011), Environmental Management Act (EMA), Zambia Environmental Management Agency (ZEMA), and Local Government Act. The generation of waste should be minimized, wherever practicable, and waste should, in order of priority, be re-used, re-cycled, recovered and disposed of safely in a manner that avoids creating adverse effects. The Act further outlines some responsibilities of people concerning the waste that they generate. It states that, a person shall not collect, transport, sort, recover, treat, store, dispose of, or otherwise manage waste in a manner that results in an adverse effect or dispose of waste in such a manner that it becomes litter (GRZ, 2011). More than 30% of the population of Lusaka have waste collection through informal service providers who are not registered with the Council and therefore do not use the controlled disposal facility (UN-Habitat, 2010). Greenhouse effect is one problem of poor waste disposal, but contamination through improper waste disposal is also problematic. Burning of waste without incinerators created air pollution and risked fire breakouts or explosions (Kasala, 2014).

Despite the legal and institutional frameworks that are in place to enhance the efficient handling and management of waste, there is still inefficient waste management in the city of Lusaka. Blessings Twavwe Namonje (2019). The inefficiency in the management of waste in many townships is evidenced by the visible waste found along the streets and in the drainages. The different types of household waste are disposed of carelessly within the urban environment. In order to maintain quality urban life, waste generated needs to be disposed of efficiently (Doan, 1998).

### Theoretical Framework

The study used the Theory of Reasoned Action (TRA) to determine the applicability in disposal of energy saving bulbs proposed by Martin Fishbein and Icek Azjen (Azjen & Fishbein, 1980). Human development is the key to sustainable development and is a continuous process of improving people’s knowledge, attitudes and practices through education, training and communication campaigns in order to increase their productivity and income while protecting the environment (Azjen & Fishbein, 1980).

The primary purpose of the TRA in the study was to understand an individual’s voluntary behavior by examining the underlying basic motivation to perform an action in disposing of fluorescent energy saving among residents in Mtendere Compound. TRA states that a person’s intention to perform a behavior is the main predictor of whether or not they actually perform that behavior. Exploring the consumer’s knowledge and attitudes and relating it to their actual practices on management and disposal of compact fluorescent light bulbs approximates the components of the theory. If a person perceives that the outcome from performing behavior is positive, he or she would have a positive attitude toward performing that behavior.

This model used as tool for analysis in the levels of knowledge and practices and could be applied to all kinds of interventions. The independent variables consisted of the respondents’ age, gender, education, and income, have been shown in past studies to affect the knowledge as well as practice of solid waste management. The dependent variables, on the other hand, included the respondents’ knowledge, attitudes and practices on solid waste management. These variables, in turn, determine, to a large extent, the kind of planning or campaign program that would be produced for ecological management of energy-saving bulbs.

## METHOD

### Study design

A cross-sectional design with a quantitative approach was used in this research study. It was a quantitative approach as questionnaires were used to collect data. It was a cross-sectional design as the study described the situation or examined that which currently existed in a population (Polit & Beck, 2012).

### Study Site

The study was conducted in Mtendere Compound a low-income, high-density settlement in the east side of Lusaka City. Zambia. Mtendere Compound, formerly known as Chainama Hills, is compound of Lusaka Zambia founded in 1967 and the name means “peace” in the native language. The compound is located about 10 Kilometers drive from the town centre of the capital of Lusaka and was electrified in 1987. The compound is characterized with over-crowding, lack of adequate safe water, poor sanitation, hygiene practices, waste management practices and poverty.

### Study Population

The study participants were drawn from among households in Mtendere compound and both women and men who are above the age of 18 years participated in the study.

### Inclusion Criteria

Adults aged 18 years and above, both female and male, who live in electrified homes in Mtendere, were included in the study.

#### 3.6 Exclusion criteria

People living in houses that were recently built or connected to electricity were excluded from the study.

#### 3.7 Sample Size

A total of Hundred and Twenty-three (193) participants (120 women and 73 men) from Mtendere were selected randomly. The total number of Hundred and Twenty-three (193) was arrived at as it was enough to generalize the findings of the research of the entire population. This was based on random sampling of the target population because all participants were given an equal chance of participating in the study. A questionnaire was then administered. The sample size was determined using Slovin’s formula to estimate the number of participants to be included in this study. This formula is suitable for estimating the minimum sampling size when nothing about a population’s behavior is known.

To arrive at the number of households that were used to derive the sample size from, 50 % of houses of Mtendere were assumed to be electrified using a 95% confidence interval and a margin error of 0.05. 385 houses were estimated to be electrified and the use Israel Table to determine the sample size.

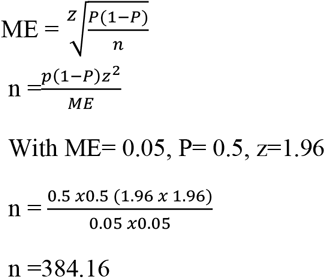

384.16 was rounded up to 385 houses and 193 participants were interviewed in the study that’s half of the houses that were assumed be electrified

### Sampling

Cluster random sampling, which is a form of sampling in which groupings or clusters where selected, typically with successive subsampling of smaller units (Polit & Beck, 2012) was used in the study. Mtendere Compound is a wide area and has a population of 120,000 people and is divided into three area or sections namely A, B, and C which were divided into three clusters. A systematic random sampling method was used to arrive at the participants’ households to participate in the study. Each household had its stand number starting point from 1 to house number 14 and the numbers were placed in a cup and the selected number was the starting point with the count of every 14^th^ number in the section. From a sampling frame for a population of 900 households in Section A, 65 households were randomly sampled. Additionally, section B had a total number of 795 households with each data collector having to stand at household number from 1 to 13, and 64 households were randomly sampled. The numbers were placed in cup to have the starting point and the selected picked number was the starting point with the count of every 13^th^ number in section B. Furthermore, section C had 855 households with each data collector having to stand at household number from 1 to 13 and 64 households were randomly sampled and the numbers were placed in cup and the selected picked number was the starting point with the count of every 13^th^ number in section C. All participants in total were 193. This sample size was determined based on the Krejcie and Morgan criteria for determining the sample size at 95 percent and 99 percent confidence levels (Krejcie & Morgan, 1970).

### Data Collection Methods

A questionnaire on energy-saving bulbs and their proper handling and disposal was used to collect data from participants (Foddy, 1994).

Each participant was interviewed at home using the structured questionnaire. Various actions were taken to ensure the quality of the questionnaire. First, all interviewers received systematic training for several days before the survey. Training provided the interviewers with an opportunity to become familiar with the survey objectives, content, and plan for implementation in the study. The training of interviewers mainly included the following areas: survey purpose; roles and responsibilities; content and use of the questionnaires; survey forms and materials; appropriate interviewing techniques, including listening skills and probing techniques. An English version of the questionnaire was developed, but interviews were translated for the field operation.

### Validity and Reliability

A small-scale pilot study was performed in Mtendere Compound on 10 households to assess the validity and reliability of the questionnaire. This was done to test if the questionnaire measures the knowledge and practices as intended in the study (Keith, 2005). Feedback was carefully reviewed with the help of senior researchers and there were no changes made to the questionnaires after the research study was conducted for validation.

### Data Analysis

Data was analysed using Statistical Package for the Social Sciences (SPSS) version 18 and descriptive Statistics were provided. Analysis is the ‘breaking up’ of something complex into smaller parts and explaining the whole in terms of properties of, and relationships between these parts. This is a necessary process for a researcher as it works as a reductionist process of the data gathered to make sense of it (Robson & McCartan 2016). The presentation of data is organized in tables, pie graphs, and bar charts that reflect frequencies and percentages of demographic data of the assessment of knowledge and disposal practices of spent and broken energy-saving bulbs.

### Dissemination of results

The results will be disseminated to the University of Zambia Library, the Zambia Environmental Management Agency (ZEMA), as well as Lusaka City, including the Ministry of Local Government. They were used be as a justification for the need to create policies that would influence the proper disposal of mercury-containing bulbs for public safety.

### Ethical Considerations

Ethics refers to rules of conduct, typically to conformity to a code or set of principles. Cohen *et al* (2011) argue that the questionnaire is always an intrusion into the life of a respondent and qualitative data analysis raises the question of identifiability, confidentiality and privacy of individuals. To that effect, the researcher had ethical obligation to fulfil the requirements such as those mentioned above. With this in mind the researcher adhered to the ethical standard practice in scientific research. Ethical approval was obtained from the University of Zambia Biomedical Research Ethics Committee (UNZABREC). Further permission was granted by the implementing institution (ACEIDHA) and the National Health Research Authority under the Ministry of Health. Verbal permission was obtained from chairman. This means the researcher was only able to carry out research, ask questions, and organise focus group discussions after explaining to the interviewees the reason of the study and the intended outcomes for both the researcher and the interviewees. Each household was informed that their participation in the exercise was voluntary and that the purpose of collecting data was purely academic. Answers were kept strictly confidential and never associated with names. This was in line with Desai and Potter (2012). The researcher also sought permission to record the interviews and also to take photographs for the respondents who were willing to be recoded and photographed. In an event that the interviewees felt uncomfortable do so, the researcher abided by their decision. No payment or coercion was made to solicit participation from respondents. Respondents were mainly household members 18 years old and above.

## RESULTS

### Demographic data

The socio-demographic data for the study comprised the gender, age, employment status, marital status, and level of education of respondents.

Table 1 shows the gender of respondents: females were 120 (62%) participants and males were 73(38%) participants. The study suggests that there were more Female participants than male ones. Additionally, participants’ education levels came as follows: Those who did not go to school were 32(17%) respondents of which the questions were read to them in a local language questionnaire and those who did Primary school were 80(41%) respondents. Those who did Secondary were 55(28%) participants while those who did Tertiary were 26(13%) respondents. The employment status was as follows: The Unemployed were 69(36%) participants while the self-employed were 102(53%) participants. The employed were 22(11%) participants. The study suggests that the self-employed were the majority while employed were few.

**Table 1:** Socio-demographic characteristics of respondents.

| VARIABLE | FREQUENCY | PERCENTAGE |
|---|---|---|
| Sex |  |  |
| Male | 73 | 38% |
| Female | 120 | 62% |
| TOTAL | 193 | 100% |
| AGE |  |  |
| 18 to 30 years | 48 | 25% |
| 31 to 45years old, | 68 | 35% |
| 46 to 55 years old | 45 | 23% |
| 56 years old and above. | 32 | 17% |
| TOTAL | 193 | 100% |
| EDUCATION LEVEL |  |  |
| Did not go to school | 32 | 17% |
| Primary | 80 | 41% |
| Secondary | 55 | 28% |
| Tertiary | 26 | 13% |
| TOTAL | 193 | 100% |
| EMPLOYMENT STATUS |  |  |
| Unemployed | 69 | 36% |
| Self-employed | 102 | 53% |
| Employed | 22 | 11% |
| TOTAL | 193 | 100% |

#### 4.2 Knowledge and practice characters

This section determines the knowledge about disposal methods of energy-saving bulbs and the associated health and environmental concerns of mercury contained in energy-saving bulbs among households in the Mtendere Compound.

Table 1 show responses from questions during the research study. Respondents were asked if they had knowledge on differentiating incandescent bulbs from energy saving bulbs. Those who said ‘don’t know’ were 51(26%) participants while those who said ‘Yes’ were 23 (12%). Those who said ‘Not decided’ were119(62%) participants.

**Table 1:** Knowledge of differentiating incandescent bulbs from energy-saving bulbs.

| VARIABLES | FREQUENCY | PERCENTAGE |
|---|---|---|
| Do you know about |  |  |
| differentiating incandescent bulbs from energy-saving bulbs? |  |  |
| Don’t know | 51 | 26% |
| Yes | 23 | 12% |
| Not decided | 119 | 62% |
| TOTAL | 193 | 100% |
| What do you use to wipe down an area where broken energy-saving bulb has occurred |  |  |
| Mop | 34 | 18% |
| Towel | 16 | 8% |
| Bloom | 23 | 12% |
| Toilet Tissue | 7 | 4% |
| Cloth | 70 | 36% |
| Papers | 34 | 18% |
| Hands | 9 | 5% |
| TOTAL | 193 | (100%) |

Participants were asked materials for use to wipe down an area where broken energy saving bulb has occurred and the responses were: Mop 34(18%) participants; Towel 16(8%) participants; Bloom 23(12%) participants; Toilet Tissue 7(4%) participants; Cloth 70(36%) participants; Papers 34(18%) participants and Hands 9(5%) participants. cloth appeared to be the most used material in the study while the bare hand was the less used.

Figure 4 above shows responses from participants when they were asked if they do burn out energy-saving bulbs or not. Some 66(34%) respondents mentioned they were ‘not sure’ while 97(50%) respondents mentioned ‘yes’ and 30(16%) respondents mentioned ‘no’. The study suggests that those who mentioned yes were the majority while those who mentioned ‘no’ were the few.

**Figure 4.**
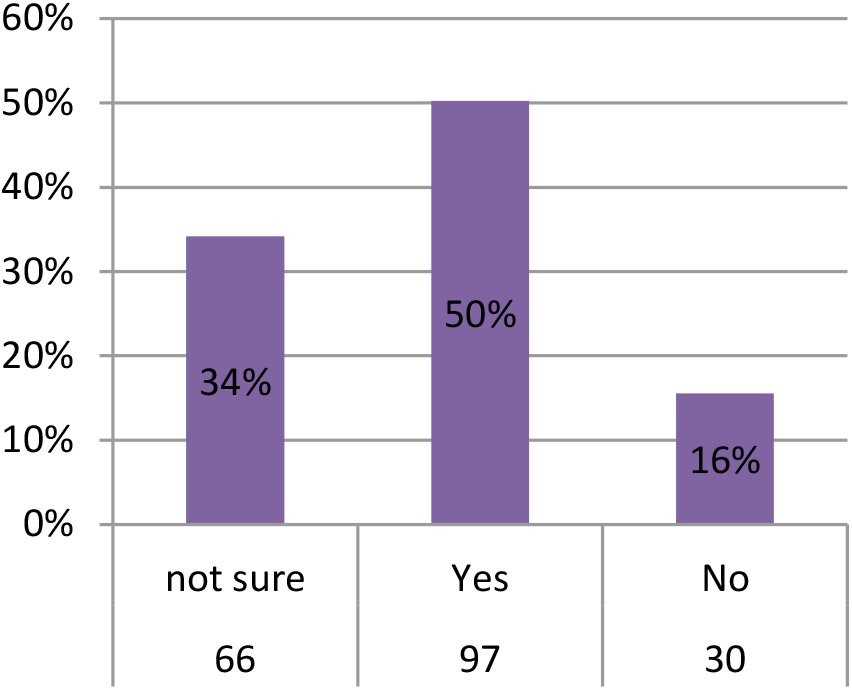
Burnt out energy-saving bulb N= 193

Figure 5.: above shows if the participants have ever discarded any energy saving bulb. To this question 145(75%) participants mentioned yes, and 48(25%) participants mentioned no. The study suggests the majority discarded broken energy saving bulbs before.

**Figure 5.**
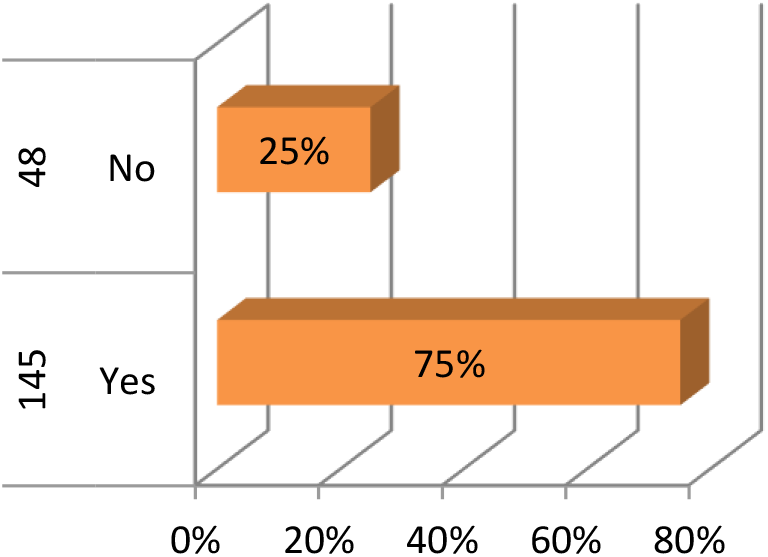
Having ever discarded any energy-saving bulb

Figure 6 above shows where participants spent energy-saving bulbs is disposed when it is broken after use and 75(39%) participants mentioned general household waste while 26(13%) participants mentioned selective-collecting stations. Some 20(10%) participants mentioned land abandoned or sidewalks, 18(9%) participants mentioned landfills, and 54(28%) participants mentioned pit latrines. The study suggests that most participants threw their broken energy-saving bulbs in general households and a few threw them on landfills.

**Figure 6.**
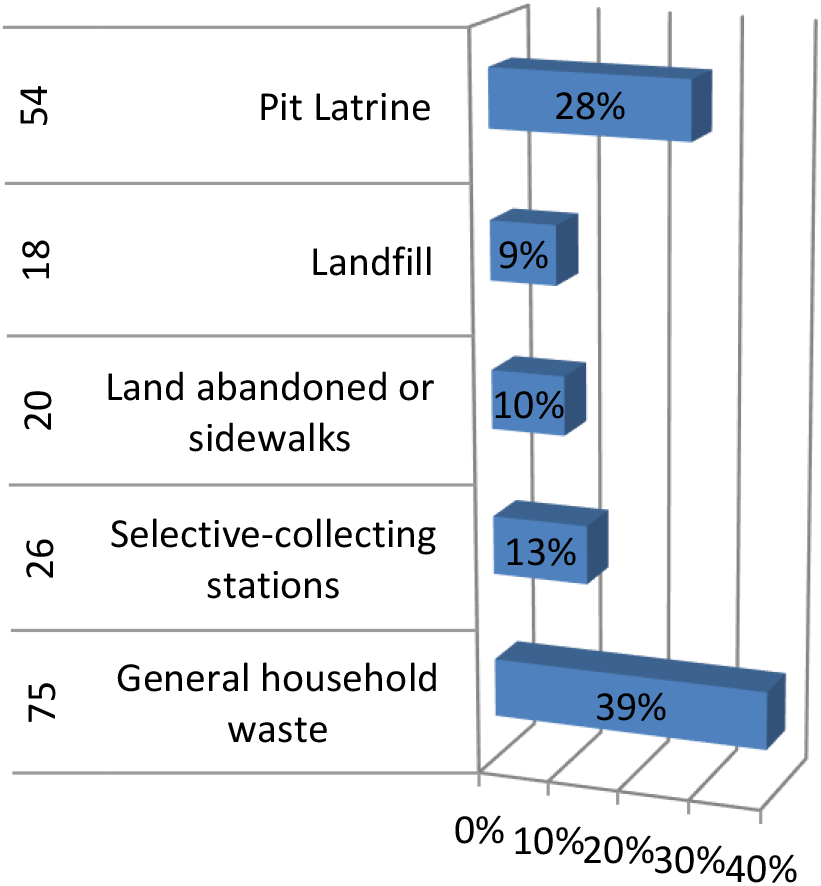
Where spent energy saving bulbs is disposed

Figure 7 above shows responses on knowledge of participants if mercury being one of the components of energy saving bulbs and some 39(20%) participants mentioned they are ‘not sure’, 58(30%) participants mentioned yes, and 96(50%) participants mentioned no. The study suggests that the majority didn’t know that mercury is one component used in energy saving bulbs.

**Figure 7.**
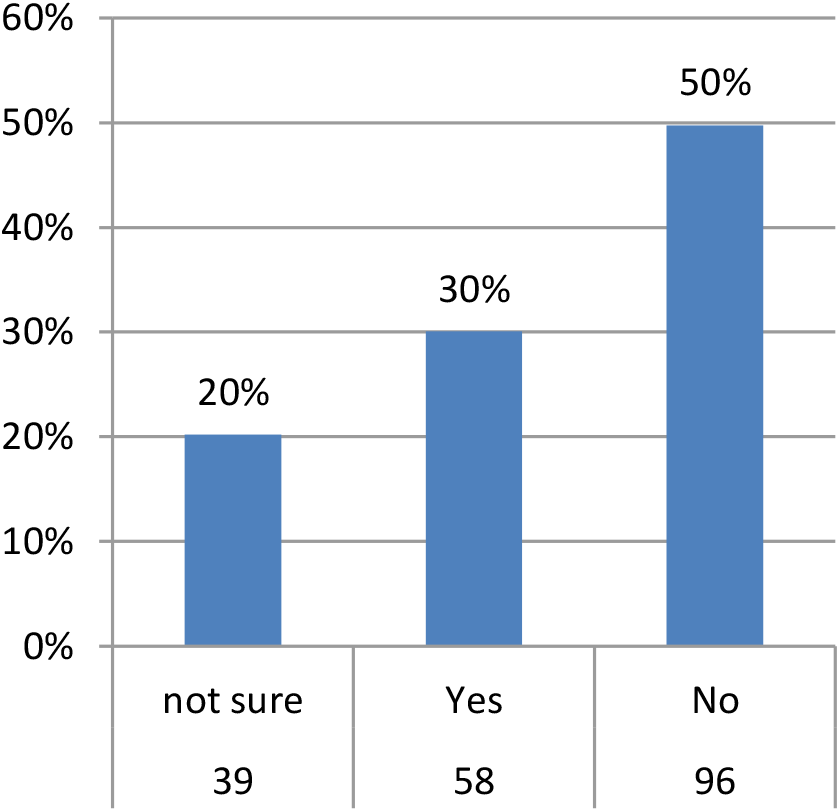
Knowledge of mercury being one of the components of energy-saving bulbs

Table 2 above shows how different participants were responding to variables during the research study. Participants were asked if they knew that mercury contaminating the environment and was dangerous to health. Those who said ‘Don’t know’ were 31(16%) participants while who said ‘Yes’ were 82(42%) participants and who said ‘Not decided’ 67(35%) participants. The study suggests that those who said YES” were the majority.

**Table 2:** Knowledge of mercury contaminating the environment and disposal practices.

| VARIABLES | FREQUENCY | PERCENTAGE |
|---|---|---|
| Do you know that mercury contaminates the environment and is dangerous to health? |  |  |
| Don’t know | 31 | 16% |
| Yes | 82 | 42% |
| Not decided | 67 | 35% |
| TOTAL | 193 | 100% |
| Do you know that disposing of debris should be in a sealable plastic bag after cleaning broken energy saving bulb |  |  |
| Never | 51 | 26% |
| Sometimes | 68 | 35% |
| Always | 61 | 32% |
| Often | 51 | 26% |
| TOTAL | 193 | 100% |

Respondents were asked if they knew how to dispose of debris in a sealable plastic bag after cleaning broken energy saving bulb. The responses were as such:

Those who said ‘Never’ were 51(26%) participants while those who said Sometimes were 68(35%) participants. Those who said ‘Always’ were 61(32%) participants while those who said Often were 51(26%) participants.

Figure 8 above shows participants having places that collect spent energy-saving bulbs in Mtendere and 64(36%) participants mentioned don’t know, 25(14%) participants mentioned yes, and 91(51%) participants mentioned no. majority suggested that they had no place that collect broken energy-saving bulbs as shown in the study.

**Figure 8.**
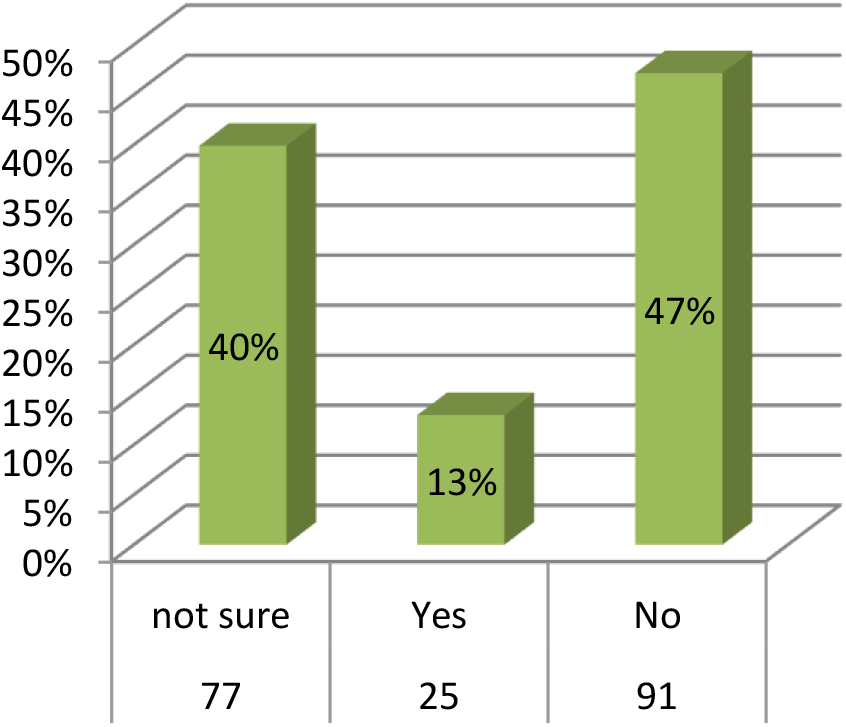
Having places that collect spent energy saving bulbs in Mtendere

Figure 9 above shows responses of participants on breaking or crushing of spent energy saving bulb before disposal and 33(17%) participants mentioned not sure’ while, 119(62% participants) mentioned yes, and 41(21%) participants mentioned no. the study suggests that the majority agrees to have broken the energy saving bulb before disposal and the minority were not sure if they did or not.

**Figure 9.**
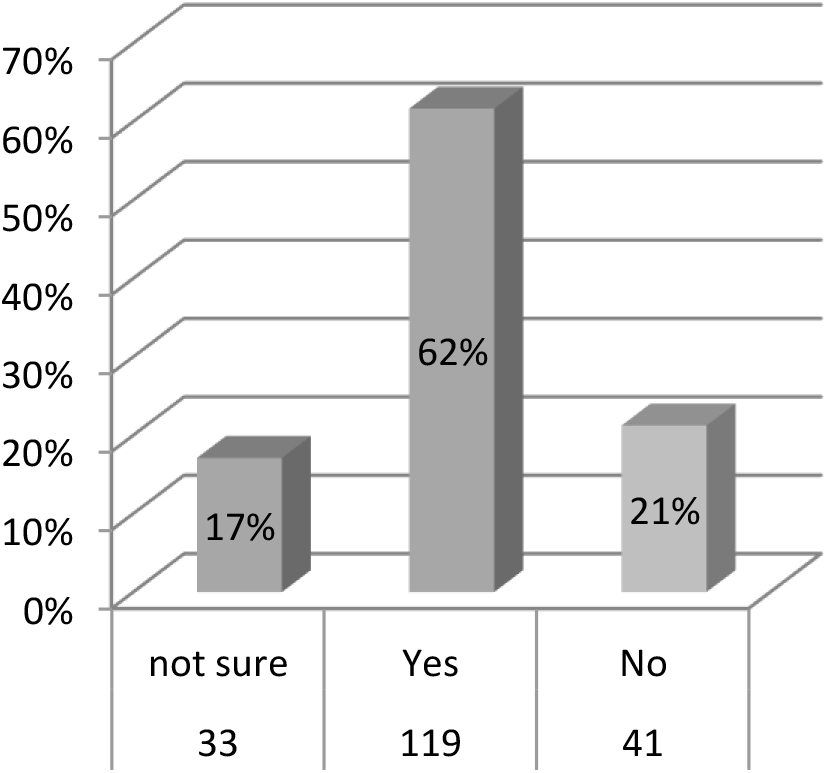
Do you crush of break the energy saving before disposal?

Figure 10 above shows if participants were getting information about the proper disposal of energy-saving bulbs, 35(18%) participants mentioned don’t know, 33(17%) participants mentioned yes, and 125(65%) mentioned no. The study suggests that the majority had no information on the disposal of energy saving bulbs.

**Figure 10.**
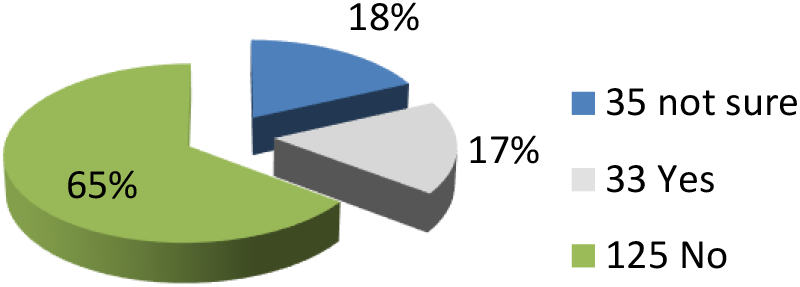
Getting information about proper disposal of energy saving bulbs

This section determines the practices regarding the handling of broken mercury-coated bulbs by Mtendere households.

Figure 11 above shows participants using rubber or latex gloves to remove broken glass of energy-saving bulbs, 95(49%) participants mentioned never, 45(23%) participants mentioned seldom, 23(12%) participants mentioned sometimes, 18(9%) participants mentioned often, and 12(6%) participants mentioned always. The study suggests that the majority never use rubber gloves to handle or remove broken energy saving bulbs.

**Figure 11.**
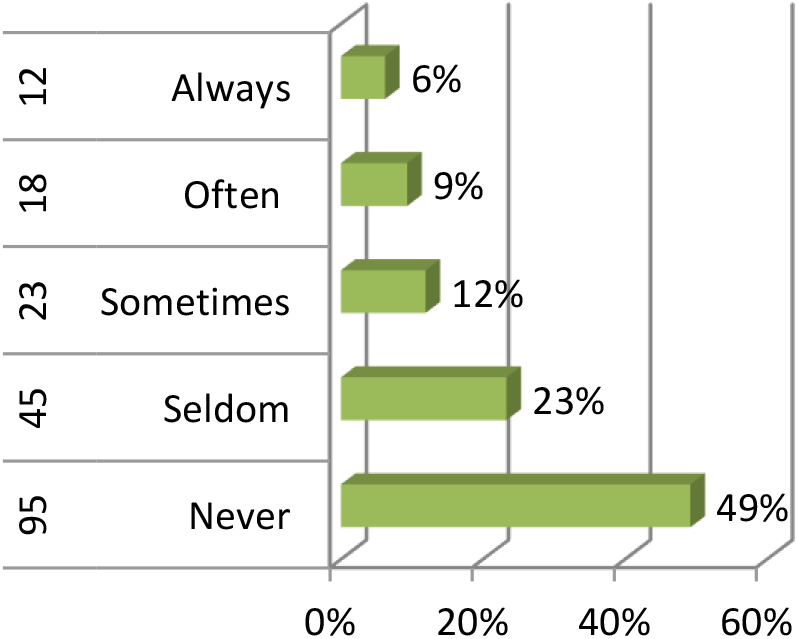
Using rubber or latex gloves to remove broken glass from energy-saving bulbs

Figure 12 above shows if participants do remove people and pets from the room in which energy saving bulb has broken, 105(54%) participants mentioned never, 19(10%) participants mentioned seldom, 37(19%) participants mentioned sometimes, 21(11%) participants mentioned often, and 11(6%) participants mentioned always. The study suggests the majority don’t remove people or pets when removing broken energy-saving bulbs.

**Figure 12.**
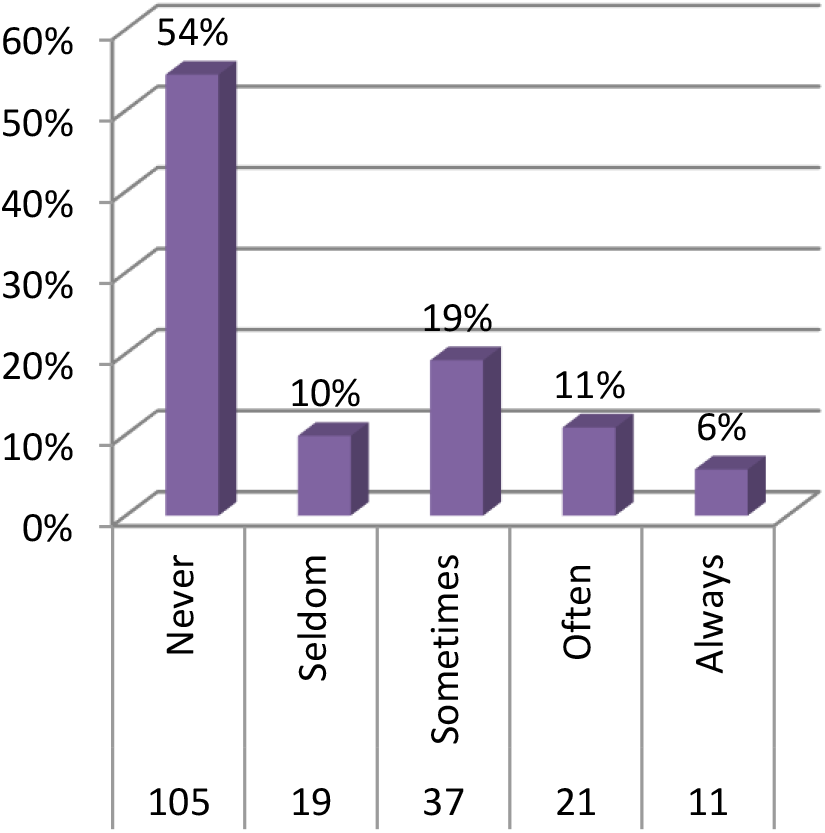
Removing people and pets from the room in which energy saving bulb has broken

Figure 13 above shows things participants used to clean up broken energy-saving bulbs, 76(35%) participants mentioned broom, 19 (10%) participants mentioned vacuum cleaners, 42 (22%) participants mentioned stiff paper, and 56 (29%) mentioned dustpan. The study suggests broom as the most commonly used item in cleaning broken energy saving bulbs.

**Figure 13.**
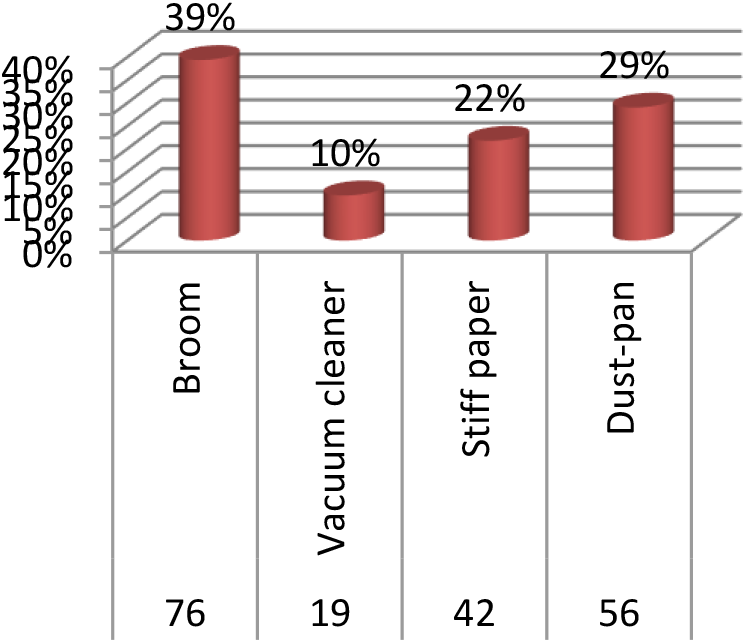
Things used to clean up broken energy-saving bulbs

Figure 14 above shows if participants kept the heater or fan turned off when an energy-saving bulb broke and 93(48%) participants mentioned never, 26 (13%) participants mentioned seldom, 32 (17%) participants mentioned sometimes, 31 (16%) participants mentioned often, and 11 (6%) participants mentioned always. The majority never turned off the heater or fan during broken energy bulb removal.

**Figure 14.**
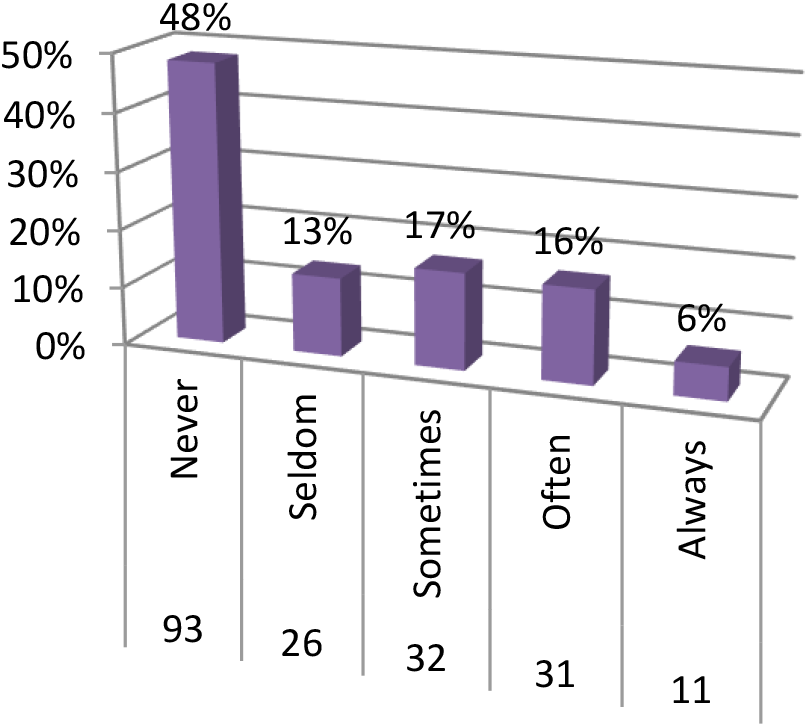
Keeping the heater or fan turned off when an energy-saving bulb has broken

Figure 15 above shows participants practices such as washing their faces, hands and changing clothes after cleaning broken energy-saving bulbs and 83(43%) participants mentioned never, 33(17%) participants mentioned seldom, 39(20%) participants mentioned sometimes, 23(12%) participants mentioned often, and 15(8%) participants mentioned always. The majority don’t wash their face and hands after handling broken spent energy bulbs as shown in the study.

**Figure 15.**
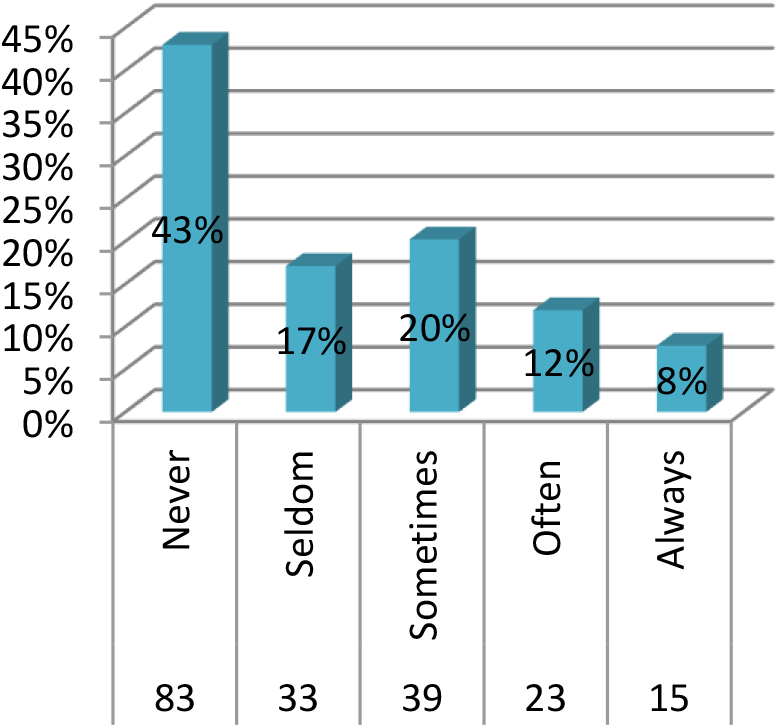
Below shows if participants their wash faces and hands after handling energy-saving bulb

## DISCUSSION

### Introduction

This chapter provides a comprehensive analysis of the results gathered from evaluating households in Mtendere Compound, Zambia, on their knowledge and disposal methods for used and damaged energy-saving bulbs. The discussion combines the findings of the study with its objectives, theoretical frameworks, and relevant literature. The goal is to comprehend the importance of the results, discuss their ramifications, and give a complete picture of the state of household knowledge and practices concerning the safe handling and disposal of energy-saving bulbs, which frequently include toxic materials like mercury. This chapter also points out the variables that affect knowledge levels and disposal behaviors, highlights shortcomings in public knowledge, and explores possible measures required to encourage safe and environmentally friendly practices. The conversation lays the groundwork for suggestions and policy implications found in the following chapter by also examining the larger public health and environmental impacts of improper disposal.

Table 4.2 reflects that most of the participants (62%) not decided in Mtendere compound clearly indicates that the majority of the residents lack knowledge about energy saving bulbs. Respondents were asked if they had knowledge on differentiating incandescent bulbs from energy saving bulbs. Those who said do not know’ were 51(26%) participants while those who said ‘Yes’ were 23 (12%). Those who said ‘Not decided’ were 19(62%) participants. Participants were asked materials for use to wipe down an area where broken energy saving bulb has occurred and the responses were: Mop 34(18%) participants; Towel 16(8%) participants; Bloom 23(12%) participants; Toilet Tissue 7(4%) participants; Cloth 70(36%) participants; Papers 34(18%) participants and Hands 9(5%) participants. cloth appeared to be the most used material in the study while the bare hand was the less used. According to Minnesota Protection Control Agency (2007), mercury coated bulbs must be handled and disposed properly to avoid polluting the environment and posing a health threat. The agency further indicate that mercury coated bulbs may not be disposed in a landfill, streams and yards, but must be recycled. Eco-South Travel (2017) indicates that most people who use mercury coated bulbs are unaware that each bulb contains between 5 and 30mg of mercury and exposure to improperly disposed broken bulbs could lead to adverse health effects.

Results on Figure 4 shows responses from participants when they were asked if they do burn out energy saving bulbs or not in Mtendere compound. Moreover, 66(34%) respondents mentioned they were ‘not sure’ while 97(50%) respondents mentioned ‘yes’ and 30(16%) respondents mentioned ‘no’. The study suggests that those who mentioned yes were the majority while those who mentioned ‘no’ were the few. These results were similar to a study conducted by Mauro, Jarbas, Gilmar & Mamerto (2011) to investigate consumer practice on the disposal of fused mercury coated bulbs in Sao Paulo, Brazil. The results indicate that about 100 million mercury coated bulbs are used a year, and only 6% is recycled. The study concludes that most of the bulbs are disposed improperly (Mauro et al., 2011).

Figure 5 shows if the participants have ever discarded any energy saving bulb. To this question 145(75%) participants mentioned yes, and 48(25%) participants mentioned no. The study suggests the majority discarded broken energy saving bulbs before. Furthermore, Figure 6 shows where participants spent energy saving bulbs is disposed when it is broken after use and 75(39%) participants mentioned general household waste while 26(13%) participants mentioned selective-collecting stations. Moreover, 20(10%) participants mentioned land abandoned or sidewalks and 18(9%) participants mentioned landfill, and 54(28%) participants mentioned pit latrine. The study suggests that most participants throw their broken energy saving bulbs in general household and a few threw them on land fill. The result concludes that lack of knowledge about proper disposal methods of mercury coated bulbs leads to improper handling of broken mercury coated bulbs.

Figure 7 above shows responses on knowledge of participants if mercury being one of the components of energy saving bulbs and some 39(20%) participants mentioned they are ‘not sure’, 58(30%) participants mentioned yes, and 96(50%) participants mentioned no. The study suggests that the majority did not know that mercury is one component used in energy saving bulbs. According to Minnesota Protection Control Agency (2007), mercury coated bulbs must be handled and disposed properly to avoid polluting the environment and posing a health threat. The agency further indicate that mercury coated bulbs may not be disposed in a landfill, streams and yards, but must be recycled. Furthermore, Table 3 shows how different participants were responding to variables during the research study. Participants were asked if they knew that mercury contaminating the environment and was dangerous to health. Those who said ‘Don’t know’ were 31(16%) participants while who said ‘Yes’ were 82(42%) participants and who said ‘Not decided’ 67(35%) participants. The study suggests that those who said “YES” were the majority. Respondents were asked if they knew how to dispose of debris in a sealable plastic bag after cleaning broken energy saving bulb. The responses were as such: Those who said ‘Never’ were 51(26%) participants while those who said Sometimes were 68(35%) participants. Those who said ‘Always’ were 61(32%) participants while those who said Often were 51(26%) participants.

Figure 9 shows responses of participants on breaking or crushing of spent energy saving bulb before disposal and 33(17%) participants mentioned ‘not sure’ while, 119(62% participants) mentioned yes, and 41(21%) participants mentioned no. the study suggests that the majority agrees to have broken the energy saving bulb before disposal and the minority were not sure if they did or not. Additionally, Figure 10 shows if participants were getting information about proper disposal of energy saving bulbs and 35(18%) participants mentioned don’t know, 33(17%) participants mentioned yes, and 125(65%) mentioned no. The study suggests that the majorly had no information on the disposal of energy saving bulbs. These results were similar to a study conducted by Mauro, Jarbas, Gilmar & Mamerto (2011) to investigate consumer practice on the disposal of fused mercury coated bulbs in Sao Paulo, Brazil. The results indicate that about 100 million mercury coated bulbs are used a year, and only 6% is properly disposed. The study concludes that most of the bulbs are disposed improperly (Mauro et al., 2017).

According to the guidelines of cleaning a broken mercury coated bulb by Minnesota Protection Control Agency (2018), a disposable paper towel should be used to wipe down powder residue of broken mercury. A mop should not be used, as it is not disposed after cleaning, and will spread further mercury beads to uncontaminated areas. However, figure 11 indicates that the majority of the households (49%) in Mtendere compound use a mop to wipe down mercury coated bulbs and only a small number of the household (6%) used disposable paper towels. The reason for them to use the mop could be that a mop can be used for many months without buying a new one. Disposable towels may be expensive for them as they may not last for many days and many people have no steady income. Additionally, Figure 12 shows if participants do remove people and pets from the room in which energy saving bulb has broken and 105(54%) participants mentioned never, 19(10%) participants mentioned seldom, 37(19%) participants mentioned sometimes, 21(11%) participants mentioned often, and 11(6%) participants mentioned always. The study suggests the majority don’t remove people or pets when removing broken energy saving bulbs in Mtendere compound. Figure 13 shows materials that participants used to clean up broken energy saving bulbs and 76(35%) participants mentioned broom, 19 (10%) participants mentioned vacuum cleaner, 42 (22%) participants mentioned stiff paper, and 56 (29%) mentioned dust-pan. The study suggests broom as the most common used item in cleaning broken energy saving bulbs. Additionally, Figure 14 shows if participants keeping the heater or fan turned off when an energy saving bulb has broken and 93(48%) participants mentioned never, 26 (13%) participants mentioned seldom, 32 (17%) participants mentioned sometimes, 31 (16%) participants mentioned often, and 11 (6%) participants mentioned always in Mtendere compound. The majority never turned off heater or fan during broken energy bulb removal. Most participants in the study used a broom to sweep an area where an Energy saving bulbs had broken instead of using a stiff paper and wiped with a mob instead of with a disposable paper towel. These sweeping and wiping practices break the mercury from broken Energy saving bulbs into droplets and spread them over the area increasing the risk of exposure to people in the household (United States Environmental Protection Agency, 2019). The results of the study suggest that people in Mtendere compound generally dispose Energy saving bulbs improperly.

### Implications

The results of this research have important environmental, public health, and policy repercussions for Zambia, especially for heavily populated peri-urban areas like Mtendere Compound. Many families continue to be at risk for both acute and chronic health issues such as respiratory issues, neurological impairments, and developmental delays in children due to the widespread ignorance of the dangers of mercury exposure from energy-saving bulbs (WHO, 2022; Clarkson & Magos, 2006). The risk of mercury vapor being released into enclosed residential areas, where ventilation is usually inadequate and exposure for family members has increased and further increased by the intentional or accidental breaking of bulbs before disposal (UNEP, 2023).

From an environmental standpoint, the improper disposal of energy-saving bulbs leads to the contamination of landfills, surface water, and groundwater with mercury and other heavy metals (Li et al., 2019). Through bioaccumulation and bio magnification processes (Boening, 2000), this poses long-term ecological hazards by impacting soil quality, aquatic organisms, and food safety. If the cumulative environmental impact is not dealt with, it may exacerbate Zambia’s already weak waste management system, making it more difficult for the country to achieve its Sustainable Development Goals (SDGs), especially those related to good health and well-being, clean water and sanitation, and responsible consumption and production (UNDP, 2023).

The study’s results also reveal significant institutional and policy shortcomings. Enforcement at the national and local levels is still weak, even with current legal frameworks like the Environmental Management Act No. 12 of 2011, partly because of limited technical capacity, insufficient funding, and poor public engagement methods (ZEMA, 2022). The lack of established collection sites for hazardous household waste indicates an immediate requirement for infrastructural development, capacity enhancement, and improved inter-sectoral cooperation among important actors, such as the private sector, public health authorities, environmental agencies, and municipal councils (Chileshe & Banda, 2020).

Additionally, the need to create more organized, science-based community education programs is highlighted by the reliance on informal information channels like television and radio for risk communication. When residents are provided with accurate information and accessible disposal options, compliance with safe waste disposal practices markedly increases, according to evidence from other developing nations (Zhang et al., 2018). Without proactive measures, the issue may worsen as Zambia’s urbanizing population increasingly adopts energy-efficient lighting technologies. (Ibid)

In summary, the findings of this research have broader implications than just Mtendere Compound; they highlight systemic issues with hazardous waste management in a variety of developing nation contexts. Urgent action is needed to avert additional health risks, environmental degradation, and policy breakdowns. The results provide a basis for enhanced regulatory frameworks that incorporate community participation in hazardous waste management, better waste management infrastructure, and focused public health interventions.

## CONCLUSION AND RECOMMENDATION

### Conclusion

This research evaluated Mtendere Compound, Zambia, households understanding and disposal behaviors regarding used and damaged energy-saving bulbs. The results clearly show a significant disconnect between the community’s awareness of the possible health and environmental risks of energy-saving bulbs, especially those associated with mercury contamination, and their widespread usage. Although the majority of families recognize the financial and energy-saving benefits of these bulbs, their disposal methods are still relatively hazardous, with most people throwing used bulbs away with regular household waste.

Practices that seriously endanger both human health and the environment are caused by inadequate regulatory enforcement, limited public awareness, and a shortage of suitable disposal facilities. This scenario highlights the pressing necessity for focused interventions, such as community awareness, capacity building, and the creation of safe, accessible disposal systems. Furthermore, the environmentally responsible management of energy-saving bulbs requires improved policy frameworks that are backed by efficient monitoring and enforcement procedures.

Tackling these issues necessitates a multi-stakeholder strategy that includes the populace, manufacturers, environmental groups, and governmental bodies. Zambia can only safeguard public health and encourage sustainable environmental management in accordance with global best practices by working together to lessen the dangers of improperly disposing of energy-saving bulbs.

### Recommendations

The results of this study clearly demonstrate the pressing need to enhance public awareness and education on the proper handling and disposal of spent and damaged energy-saving bulbs in Mtendere Compound. Many families are unaware of the dangerous elements of these bulbs, especially the presence of mercury, which, if not handled properly, can pose serious health and environmental hazards (UNEP, 2012). As a result, residents should be educated about the risks of improper disposal and the necessity of adopting safe disposal practices through focused public awareness initiatives that make use of local media, community outreach programs, schools, and healthcare institutions (Ongondo et al., 2011).

In addition, the Zambian government should establish and implement explicit regulations for the collection, storage, and disposal of used and damaged energy-saving bulbs through the appropriate regulatory agencies, such as the Zambia Environmental Management Agency (ZEMA). The lack of designated disposal sites and collection locations currently contributes to improper disposal in general waste streams, resulting in environmental pollution (Puckett et al., 2002). Households may be encouraged to take an active role in safe disposal programs by establishing conveniently located collection centers within the compound and providing incentives like small financial rewards or discounts on new bulb purchases (Kahhat et al., 2008).

Furthermore, to create sustainable take-back programs and recycling facilities for energy-saving bulbs, there is a necessity for improved cooperation among government entities, private sector players (like bulb manufacturers and retailers), and non-governmental organizations (NGOs). Extended Producer Responsibility (EPR) regulations could be implemented to hold producers accountable for the management of their goods at the end of their lifespan (Manomaivibool, 2009). This strategy would not only lessen the environmental impact but also generate employment in the recycling industry.

In addition, to avoid occupational exposure and secondary contamination during waste collection and processing, waste handlers and municipal personnel should be trained on the appropriate handling of hazardous waste, including broken energy-saving bulbs (Li et al., 2012). By integrating such training into current waste management procedures, it will be possible to ensure that the entire disposal process is carried out securely and efficiently.

Finally, interventions that have been put in place should be evaluated and monitored on a regular basis to determine how effective they are. This will aid in pinpointing places that need more enhancement and make sure that safe disposal methods and knowledge sharing remain a continuous focus in Mtendere Compound (Wilson et al., 2009).

## Data Availability

Availability of data and materials
The datasets and/or analyzed during the current study are available from the corresponding author on reasonable request.

https://www.unza.zm/library

## Authors’ contributions

The author made substantial contributions to the conception and design, acquisition of data, or analysis and interpretation of data, took part in drafting the article or revising it critically for important intellectual content, gave final approval of the version to be published, and agreed to be accountable for all aspects of this manuscript.

## Funding

This manuscript research did not receive any specific grant from funding agencies in the public, commercial, or not-for-profit sectors.

## Availability of data and materials

The datasets and/or analyzed during the current study are available from the corresponding author on reasonable request.

## Ethics approval and consent to participate

Ethical clearance to conduct this study was sought from the Research Ethics and Science (RES) committee.

## Consent for publication

Written informed consent for the publication was obtained from the participants.

## Competing interests

The author declare that they have no known competing financial interests or personal relationships that could have appeared to influence the work reported in this paper.

## Acknowledgements

I am grateful to all respondents who participated in this study. Thanks are also given to the anonymous referees for any comments and suggestions that helped to produce the manuscript in its current form.

## Notes

### Competing Interest Statement

The authors have declared no competing interest.

### Author Declarations

Ethical approval was obtained from the University of Zambia Biomedical Research Ethics Committee (UNZABREC). Further permission was granted by the implementing institution (ACEIDHA) and the National Health Research Authority under the Ministry of Health.

### Summary of Updates

Yes it has been .

